# Covid-19 positive test cycle threshold trends predict covid-19 mortality in Rhode Island

**DOI:** 10.1101/2021.01.26.21250557

**Authors:** Andrew G. Bostom, Todd Kenyon, Charles B. Eaton

## Abstract

The cycle thresholds (Cts) at which reverse transcriptase polymerase chain reaction (rtPCR) tests for covid-19 become positive are intimately associated with both viral load, and covid-19 infectiousness (i.e., ability to culture live virus). Clinical data indicate lower Cts—and hence larger viral loads—independently predict greater covid-19 mortality when patients are hospitalized for symptomatic covid-19 pneumonia. We merged public covid-19 mortality data from the Rhode Island Department of Health with a de-identified dataset of n=5036 positive rtPCR test Cts from the Rhode Island Department of Health State Laboratory to explore the potential relationship between positive covid-19 test Ct distribution trends, and covid-19 mortality in the state of Rhode Island, from March through early to mid-June, 2020. Mean daily covid-19 positive test Ct data were compiled, and 7-day rolling average covid-19 mortality was offset by 21-days, given the lag between infection and death. We divided the Ct data into three strata, >32, 28-32, and <28, which were operationally defined as “not infectious,” “maybe infectious,” and “infectious,” respectively. Between late March and June, mean daily Ct values rose linearly (R-squared=0.789) so that by early June, as the covid-19 pandemic ebbed in severity, all means reached the noninfectious (Ct >32) range. Most notably, this May-June trend for Cts was accompanied by a marked, steady decline in Rhode Island’s daily covid-19 mortality. Our results suggest that monitoring, and public reporting of mean population covid-19 test Cts over time is warranted to gauge the vacillations of covid-19 outbreak severity, including covid-19 mortality trends.

## Introduction

The cycle thresholds (Cts) at which reverse transcriptase polymerase chain reaction (rtPCR) tests for covid-19 become positive are intimately associated with both viral load, and covid-19 infectiousness (i.e., ability to culture live virus).^1,2^ An rtPCR covid-19 assay system developed at the Harvard University/ Massachusetts Institute of Technology Broad Institute,^3^ currently determining covid-19 “positivity” at 108 northeastern universities—including Rhode Island’s major colleges^4^—described this *exponential* relationship:^3^ *“…the Ct values correlated strongly with the logarithm of (covid-19) RNA concentration (R-squared > 0*.*99), with the observed range from Ct =12 cycles to Ct = 38 cycles corresponding to viral loads ranging from ∼1*.*9 billion copies/mL to 8 copies/mL, respectively*

Additional clinical data indicate lower Cts—and hence larger viral loads—independently predict greater covid-19 mortality when patients are hospitalized for symptomatic covid-19 pneumonia, or other manifestations of being heavily infected by the virus.^5^ Conversely, a study which recorded Cts of patients, serially, during their hospitalization for diagnosed covid-19 pneumonia, reported that increasing Cts were accompanied by decreasing disease severity (Sequential Organ Failure Assessment; SOFA) scores.^6^ We explored the potential relationship between positive covid-19 test Ct distribution trends, and covid-19 mortality in the state of Rhode Island, between March and June, 2020.

## Methods

Public covid-19 mortality data from the Rhode Island Department of Health COVID-19 Data Tracker,^7^ were merged with a de-identified dataset of n=5036 positive rtPCR test Cts obtained through an Access to Public Records Act request to the Rhode Island Department of Health State Lab (RISHL). Standard rtPCR SARS-CoV-2 assay methodology targeting the nucleocapsid genes N1 and N2 was employed by the RISHL.^8^ Mean daily covid-19 positive test Ct data were compiled, and 7-day rolling average covid-19 mortality was offset by 21-days, given the lag between infection and death. We divided the Ct data into three strata, >32, 28-32, and <28, which were operationally defined as “not infectious,” “maybe infectious,” and “infectious,” respectively, consistent with prior reports^1,2^, including a pooled analysis of six studies^1^.

## Results

Between late March and June, mean daily Ct values rose linearly (Figure 1.; R-squared=0.789) so that by early June, as the covid-19 pandemic ebbed in severity, all means reached the non-infectious (>32) range. Most notably, Figure 2. depicts how this May-June trend for Cts was accompanied by a pronounced, steady decline in Rhode Island’s daily covid-19 mortality,

**Figure 1.**
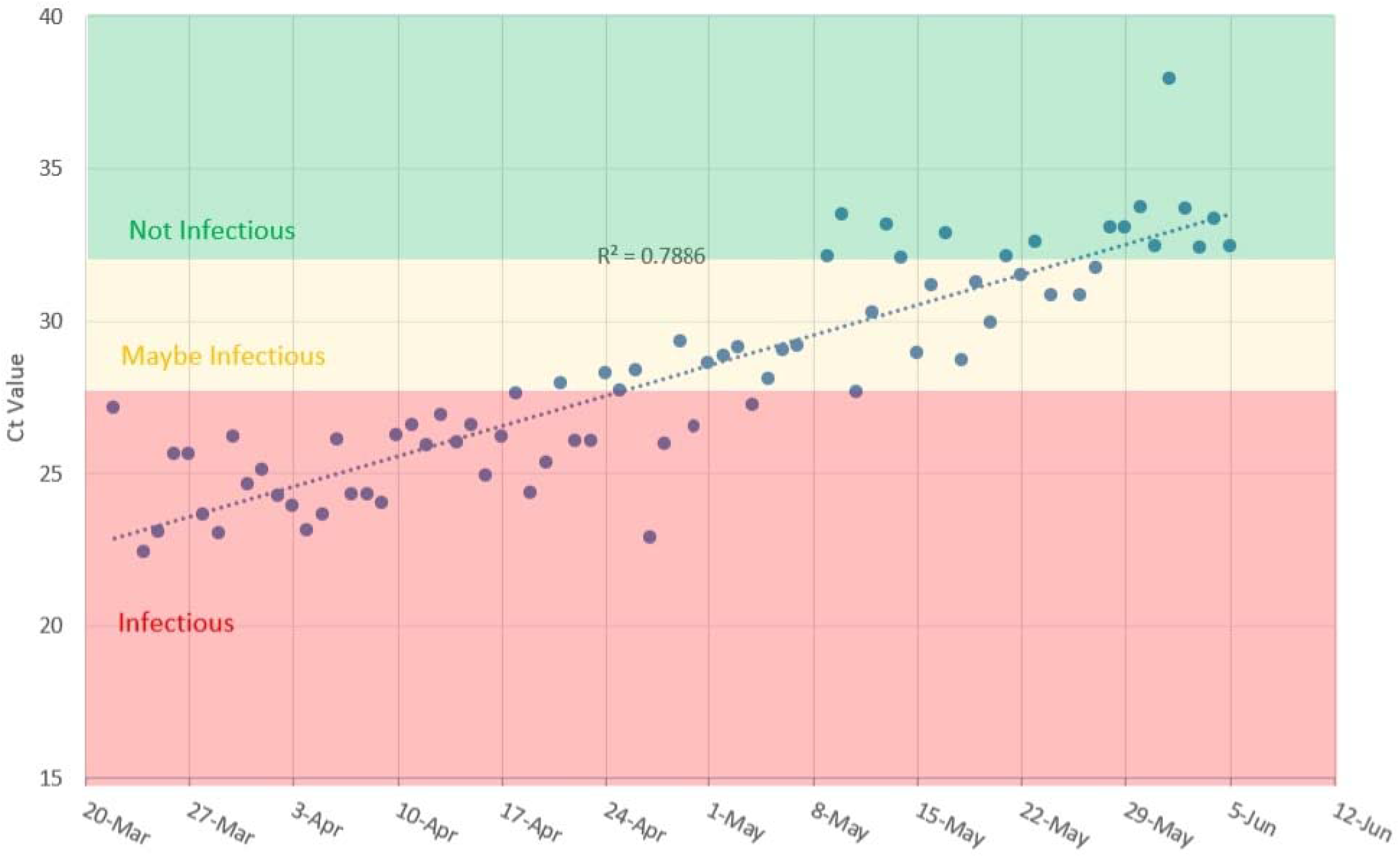
Covid-19 rtPCR^a^ mean daily Ct^b^ values, March-June, 2020. ^a^rtPCR= reverse transcriptase polymerase chain reaction ^b^Ct=cycle threshold, daily means; blue dots

**Figure 2.**
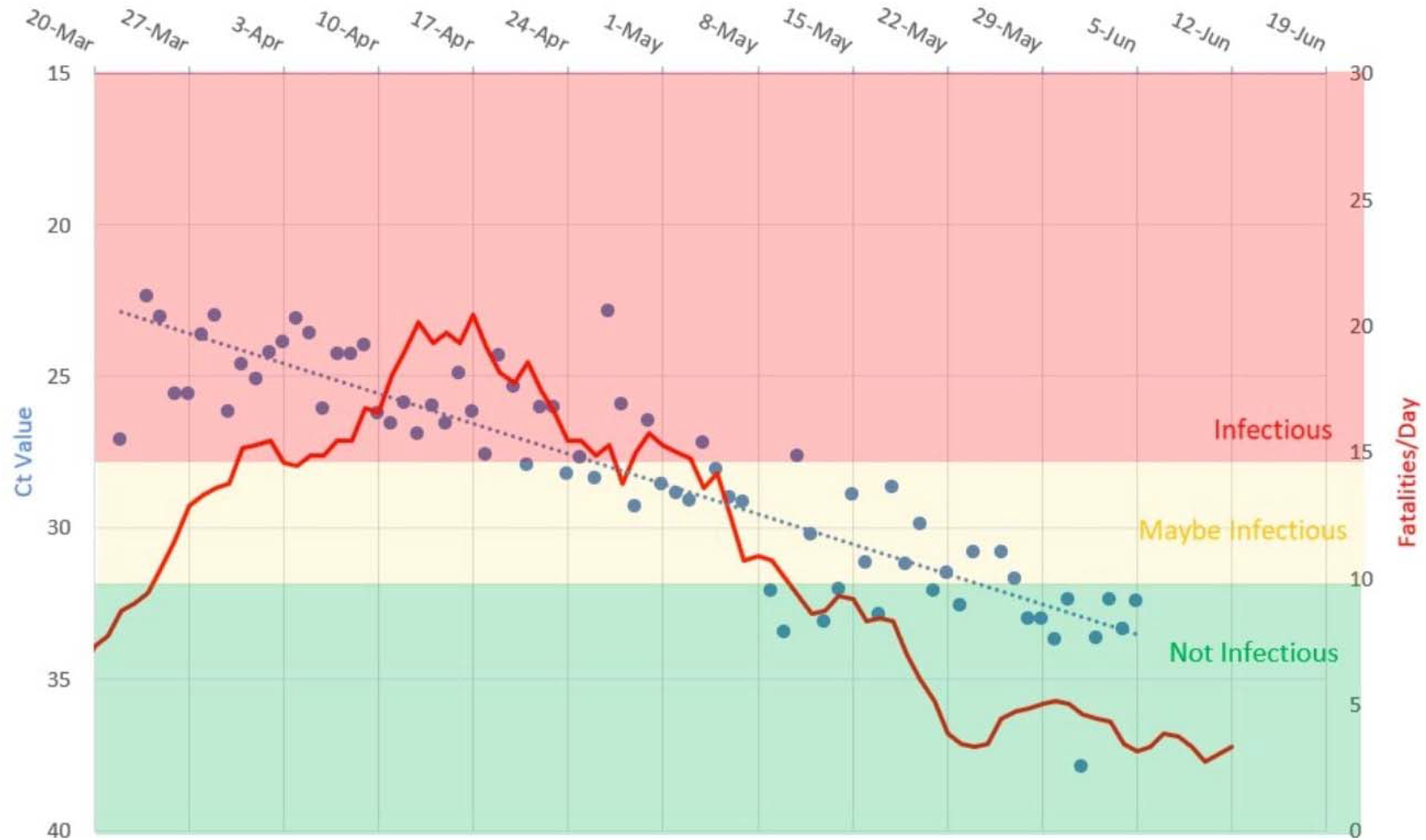
Covid-19 rtPCR positive test mean daily Ct values ^c^, and covid-19 deaths^d^. ^c^Ct=cycle threshold, daily means; blue dots ^d^21-day offset for 7-day rolling average of deaths; red line curve

## Discussion

An analysis evaluating the infectiousness of patients hospitalized with covid-19 reported that ***only*** viral loads > 10 million copies/mL, equivalent to Cts ≤ 25, were associated with isolation of infectious virus from the respiratory tract.^9^ A complementary systematic review published 12/3/30 by the Oxford University Center for Evidence-Based Medicine confirmed that covid-19 rtPCR testing patient sample Cts >30 (mean from 6-studies) are associated with an inability to culture live virus, i.e., are non-infectious.^1^

Data analyzed from the United Kingdom’s National Covid-19 Infection Survey revealed a significant impact of calendar time on the prevalence of rtPCR positives at a Ct <30: markedly fewer during mid-July to early August, versus the month of May, through mid-June, 2020.^6^ Moreover, investigators from the Bronx Montefiore Medical Center have reported lower hospital admission covid-19 rtPCR Cts were independently associated with increased covid-19 inpatient mortality.^5^ Specifically, Cts <22.9 (the lowest quartile), multivariable-adjusted for age, sex, body-mass index, hypertension, and diabetes, were associated with ∼4-fold greater covid-19 mortality risk, versus Cts >32.4 (the uppermost quartile).^5^ Using a comparable cutpoint, *a priori*, i.e., Cts >32,^2^ we externally validated these findings by demonstrating that statewide Rhode Island covid-19 mortality dropped precipitously from March to June, 2020, as mean covid-19 positive test Cts from our RISHL sample rose above 32.

## Conclusion

Our results suggest that monitoring, and public reporting of mean population covid-19 test Cts over time is warranted to gauge the vacillations of covid-19 outbreak severity, including covid-19 mortality trends.

## Data Availability

All data are publicly available

## References

1. Jefferson T, Spencer EA, Brassey J, Heneghan C. Viral cultures for COVID-19 infectious potential assessment - a systematic review. Clin Infect Dis. 2020 Dec 3:ciaa1764. doi: 10.1093/cid/ciaa1764. Epub ahead of print. PMID: 33270107.

2. Basile K, McPhie K, Carter I, Alderson S, Rahman H, Donovan L, Kumar S, Tran T, Ko D, Sivaruban T, Ngo C, Toi C, O’Sullivan MV, Sintchenko V, Chen SC, Maddocks S, Dwyer DE, Kok J. Cell-based culture of SARS-CoV-2 informs infectivity and safe de-isolation assessments during COVID-19. Clin Infect Dis. 2020 Oct 24:ciaa1579. doi: 10.1093/cid/ciaa1579. Epub ahead of print. PMID: 33098412; PMCID: PMC7665383.

3. Lennon NJ, Bhattacharyya RP, Mina MJ, Rehm HL, Hung DT, Smole S, Woolley A, Lander ES, Gabriel SB. Comparison of viral levels in individuals with or without symptoms at time of COVID-19 testing among 32,480 residents and staff of nursing homes and assisted living facilities in Massachusetts. medRxiv 2020.07.20.20157792; doi: https://doi.org/10.1101/2020.07.20.20157792

4. “Broad Institute provides COVID-19 screening for students, faculty, and staff at more than 100 colleges and universities.” September 2, 2020 https://www.broadinstitute.org/news/broad-institute-provides-covid-19-screening-students-faculty-and-staff-more-100-colleges-and

5. Choudhuri J, Carter J, Nelson R, Skalina K, Osterbur-Badhey M, Johnson A, Goldstein Y, Paroder M, Szymanski J. SARS-CoV-2 PCR cycle threshold at hospital admission associated with patient mortality. medRxiv 2020.09.16.20195941; https://doi.org/10.1101/2020.09.16.20195941

6. Zacharioudakis IM, Zervou FN, Prasad PJ, et al. Association of SARS-CoV-2 genomic load trends with clinical status in COVID-19: A retrospective analysis from an academic hospital center in New York City. PLoS One. 2020;15(11):e0242399. Published 2020 Nov 17. doi: https://journals.plos.org/plosone/article?id=10.1371/journal.pone.0242399

7. Rhode Island Department of Health COVID-19 Data Tracker https://docs.google.com/spreadsheets/d/1c2QrNMz8pIbYEKzMJL7Uh2dtThOJa2j1sSMwiDo5Gz4/edit#gid=1592746937

8. Lu X, Wang L, Sakthivel SK, et al. US CDC Real-Time Reverse Transcription PCR Panel for Detection of Severe Acute Respiratory Syndrome Coronavirus 2. Emerging Infectious Diseases. 2020;26(8):1654–1665. doi:10.3201/eid2608.201246

9. COVID-19: Shedding of infectious virus in hospitalized patients with COVID-19 Jefferson T, Spencer EA. Heneghan C. Published on August 11, 2020 https://www.cebm.net/study/shedding-of-infectious-virus-in-hospitalized-patients-with-covid-19/

10. Walker AS, Pritchard E, House T, Robotham JV, Birrell PJ, Bell I, Bell JI, Newton JN, Farrar J, Diamond I, Studley R, Hay J, Karina-Doris V, Peto T, Stoesser N, Matthews PC, Eyre DW, Pouwels KB, the COVID-19 Infection Survey team. “Viral load in community SARS-CoV-2 cases varies widely and temporally” medRxiv 2020.10.25.20219048; doi: https://doi.org/10.1101/2020.10.25.20219048

